# Perception of primary care services among the Afroperuvian population

**DOI:** 10.1101/2023.12.15.23299478

**Authors:** Elisa Juárez Chávez, Dayana Urday Fernández, Kelika A. Konda, José Villalobos Ruiz, María Sofía Cuba Fuentes

## Abstract

The Peruvian public healthcare system has various shortcomings that impact the experiences and perceptions of quality among the general population and, particularly, among minority populations, including the Afro-Peruvian community. In this context, understanding the population’s experiences in healthcare services and their perceptions regarding the treatment received from healthcare providers is crucial for developing improvement recommendations. Through in-depth interviews, our study explored the usage experiences and perceptions regarding the treatment received from public primary healthcare providers among 19 Afro-Peruvian individuals in Lima. Our analysis revealed that, in general, there is mistreatment of healthcare providers. This is not only recognized as a deficiency of the providers (human resources) but also as a consequence of a system that does not promote the quality of care, including issues such as inadequate infrastructure, low salaries, and limited time for patient care. Additionally, expressions of discrimination based on differences, primarily socioeconomic, were identified. Our findings provide insights for enhancing healthcare services in terms of treatment, effective communication, and infrastructure at the primary healthcare level.

## Introduction

The Pan American Health Organization (PAHO) has defined access to healthcare services as “the ability to use comprehensive, appropriate, timely, and quality health services when needed” (1) While there are different definitions, most concur that access implies that individuals achieve real use of services. This goal is affected by multiple factors or characteristics that influence the process of seeking and obtaining healthcare services, both inherent to the healthcare system (e.g., resources, procedures, institutions) and related to the population (e.g., perception of illness, language, cultural beliefs).

### Peruvian Health System

The Peruvian healthcare system is the set of institutions or organizations directly involved in providing healthcare services and improving the population’s health and quality of life (2). Effective access to healthcare services in Peru depends on socioeconomic status and one’s employment-status, as there are subsystems for specific population groups, each of which manages its financing and healthcare service provision processes. Therefore, it has been noted that the Peruvian healthcare system is fragmented and segmented (3). The two most important systems funded by state resources are the “Seguro Integral de Salud” (SIS), which is financed through taxes and primarily serves economically disadvantaged individuals. And EsSalud, which is funded through deductions from the salaries of individuals who are formally employed.

Regarding the complexity or resolution capacity that healthcare facilities can possess, these can be divided into three levels: primary, secondary and tertiary. Primary care serves as the entry point for the population into the healthcare system. This level is characterized by high demand with a low level of complexity since it focuses on activities related to health promotion and prevention, early diagnosis, and timely treatment, focusing on the most common health needs of individuals, families, and communities (4). Secondary care complements the care initiated at the preceding level, adding more specialization in human resources and technology (5). The third level of care represents the highest specialization and resolution capacity. Each level has human resources and technology to provide the care required. (See Table 1).

**Table 1.**
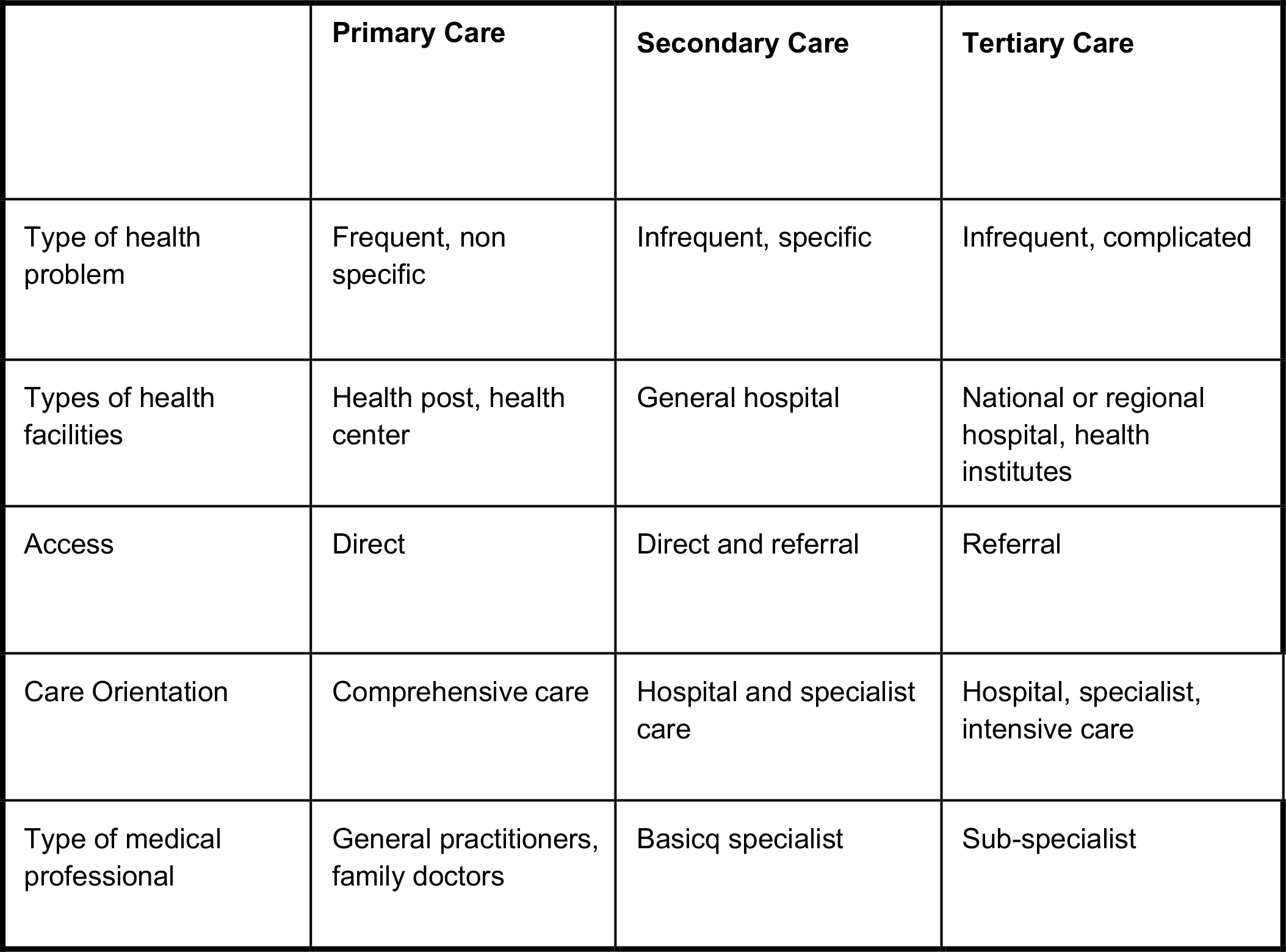
Characteristics of the Levels of Care in the Peruvian Healthcare System.

|  | **Primary Care** | **Secondary Care** | **Tertiary Care** |
| --- | --- | --- | --- |
| Type of health problem | Frequent, non specific | Infrequent, specific | Infrequent, complicated |
| Types of health facilities | Health post, health center | General hospital | National or regional hospital, health institutes |
| Access | Direct | Direct and referral | Referral |
| Care Orientation | Comprehensive care | Hospital and specialist care | Hospital, specialist, intensive care |
| Type of medical professional | General practitioners, family doctors | Basicq specialist | Sub-specialist |

Adapted from Starfield B. Primary Care: Balancing Health Needs, Services and Technology, Oxford Press New York 1998 (6)

### Access to health care in Perú

Previous studies provide evidence that the characteristics of a healthcare facility, including its infrastructure and the human resources providing services, are determining factors that influence decisions regarding the use of a healthcare establishment. In this regard, the staff’s demeanor becomes a key element in the perception of service quality and, consequently, in an individual’s future use of these services. With that said, evidence demonstrates that interactions often involve expressions of discrimination of various kinds, including racial discrimination, discrimination based on socio-economic factors, and discrimination related to the population’s level of information about how to take care of their health, among others (7) (8).

In Perú, difficulties in accessing services have led to increased use of pharmacies for consultation in the presence of uncomplicated symptoms or symptoms perceived to be uncomplicated. A recent report from the Peruvian National Institute of Statistics and Informatics (INEI) indicates that in the last quarter of 2022, 43.5% of the population with a health issue sought health care. Among them, 48.5% sought care at a pharmacy or drugstore, 30.4% went to Ministry of Health (MINSA) facilities, 12.5% visited a private clinic, and 8.9% sought care at facilities of the Social Health Insurance (EsSalud). (9).

Furthermore, access to healthcare services is not the same across the country. According to estimates from the Ministry of Culture (10) and INEI (11), access to services in the Afro-Peruvian population has improved in recent decades (increasing from 41% to over 70% between 2004 and 2017 in terms of coverage of both public and private insurance), there are significant differences in access to healthcare services between the Afro-Peruvian population and the general population or populations with different ethnic identities. For example, in 2011, a report from the Office of the Ombudsman revealed that the Afro-Peruvian population faces difficulties in accessing healthcare services, such as discrimination, differential treatment, and infrastructure barriers to access, leading to a significant percentage of Afro-Peruvians choosing not to seek healthcare services when they have an illness or emergency (12).

Health inequities result from various social determinants, making attributing them to a single cause impossible. However, it is necessary to identify specific points to improve the effective utilization of services and their quality. Qualitative methods allow for a deep exploration of the experiences of use and perceptions regarding the care provided at state-run primary-level healthcare facilities, with the expectation of addressing these aspects.

## Methods

### Study Participants

Nineteen in-depth interviews were conducted with individuals self-identifying as Afro-Peruvian, residing in metropolitan Lima, who have utilized state-run healthcare services (specifically SIS or EsSalud) at the primary care level. The fieldwork was carried out between January and April 2023. Thirteen interviews were conducted face-to-face, while 6 were conducted by telephone due to the impossibility of the participants meeting with us in person.

**Table 2.**
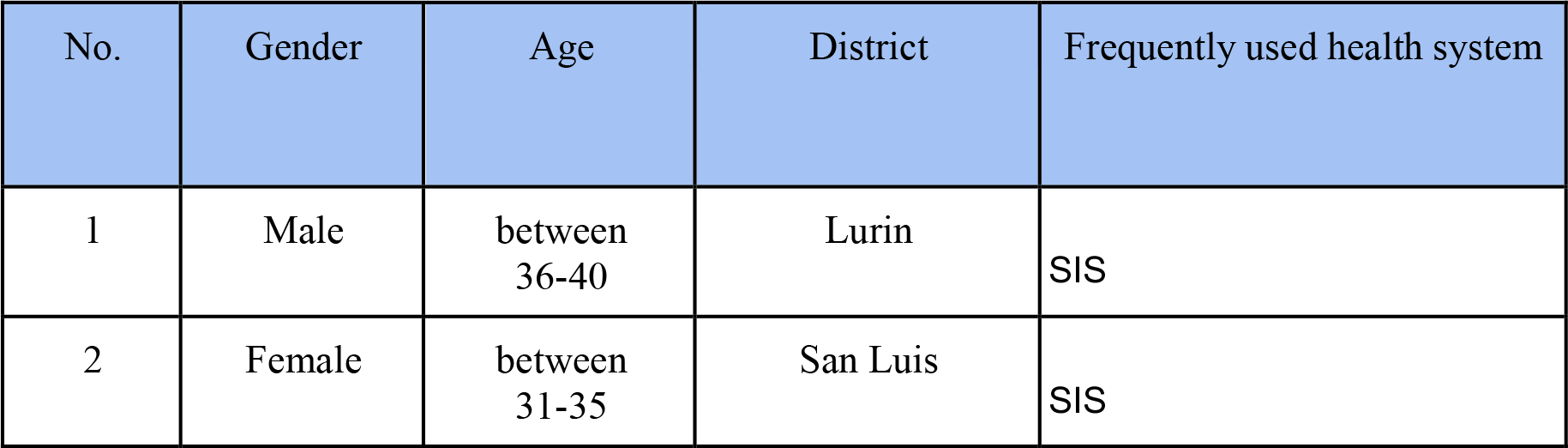
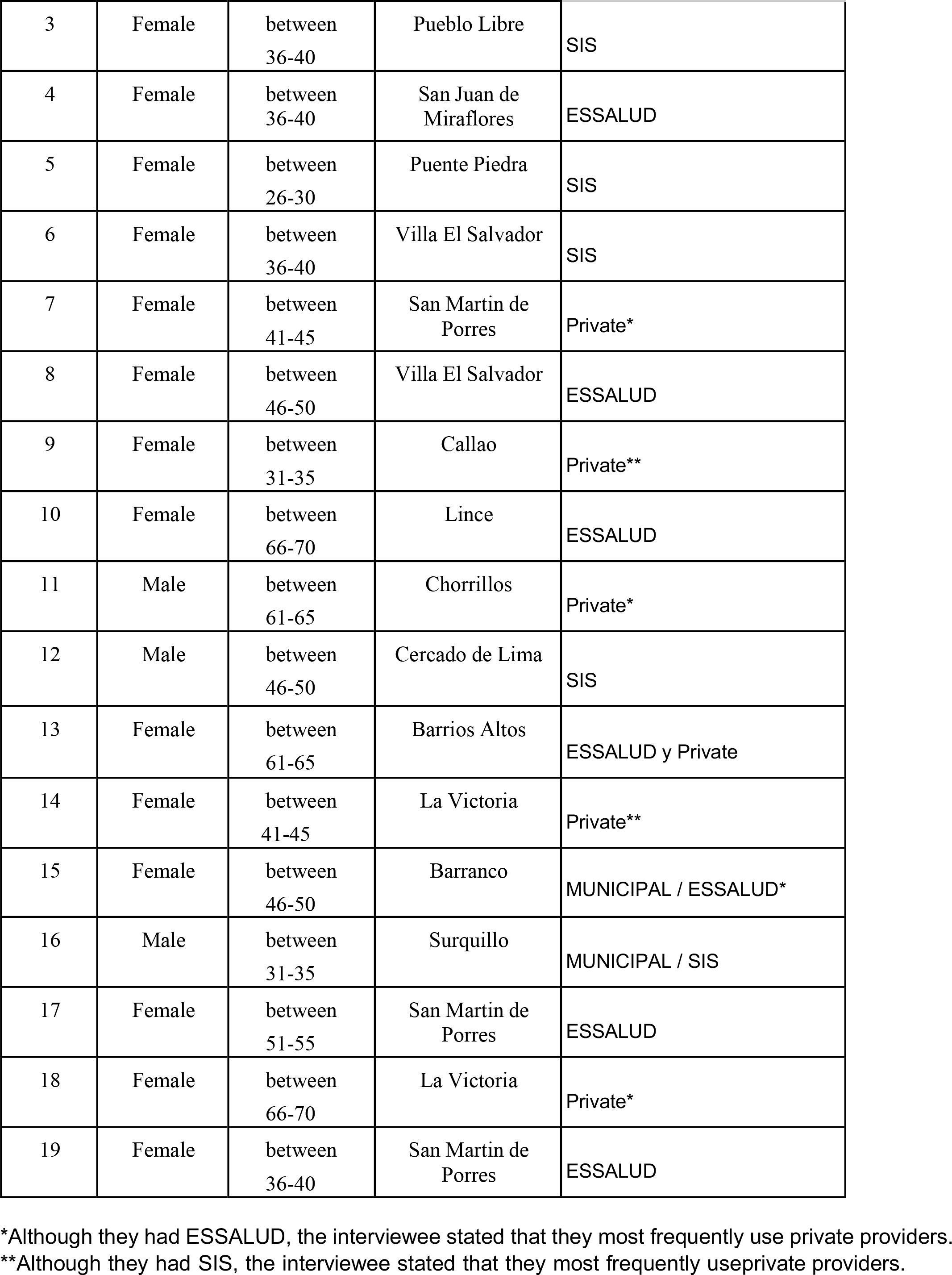
List of interviewees with their main characteristics.

| No. | Gender | Age | District | Frequently used health system |
| --- | --- | --- | --- | --- |
| 1 | Male | between 36-40 | Lurin | SIS |
| 2 | Female | between 31-35 | San Luis | SIS |
| 3 | Female | between<br>36-40 | Pueblo Libre | SIS |
| 4 | Female | between<br>36-40 | San Juan de<br>Miraflores | ESSALUD |
| 5 | Female | between<br>26-30 | Puente Piedra | SIS |
| 6 | Female | between<br>36-40 | Villa El Salvador | SIS |
| 7 | Female | between<br>41-45 | San Martin de<br>Porres | Private* |
| 8 | Female | between<br>46-50 | Villa El Salvador | ESSALUD |
| 9 | Female | between<br>31-35 | Callao | Private** |
| 10 | Female | between<br>66-70 | Lince | ESSALUD |
| 11 | Male | between<br>61-65 | Chorrillos | Private* |
| 12 | Male | between<br>46-50 | Cercado de Lima | SIS |
| 13 | Female | between<br>61-65 | Barrios Altos | ESSALUD y Private |
| 14 | Female | between<br>41-45 | La Victoria | Private** |
| 15 | Female | between<br>46-50 | Barranco | MUNICIPAL / ESSALUD* |
| 16 | Male | between<br>31-35 | Surquillo | MUNICIPAL / SIS |
| 17 | Female | between<br>51-55 | San Martin de<br>Porres | ESSALUD |
| 18 | Female | between<br>66-70 | La Victoria | Private* |
| 19 | Female | between<br>36-40 | San Martin de<br>Porres | ESSALUD |

\*Although they had ESSALUD, the interviewee stated that they most frequently use private providers.
\*\*Although they had SIS, the interviewee stated that they most frequently use private providers.

### Qualitative Interviews

The interviews took approximately 45 minutes to one hour and were conducted by an anthropologist with extensive qualitative research experience. When the interviews were done in person, compensation was provided to cover transportation expenses, and a snack was provided. In those cases when they were not in person we could not provide that compensation.

A guide containing open-ended questions focused on understanding experiences with healthcare services and their adaptation to the cultural needs of the target population, including barriers and facilitators to accessing healthcare services and the incorporation of cultural characteristics of the Afro-Peruvian population within these healthcare services, was used for the interviews. Also, we explored the experiences lived in the health facilities and perceptions about the interaction with health providers However, in all cases, the conversation centered on the interviewee’s experiences with the health services and their perception of the treatment. Even when participants reported both private and public services as the most used ones, we focused interviews on experiences and perceptions associated with public services.

### Analysis

The analysis team was composed of two of the authors of this article (First and second author). Prior to analysis, the audio files of the interviews were transcribed verbatim. Together, we constructed and discussed an initial version of the codebook. We then analyzed one of the transcriptions to assess the codebook’s utility. Subsequently, the coding of the remaining transcriptions was conducted by the second author.

Topic coding was used in this analysis. Our first set of codes was based on elements included in the interview guide. Furthermore, a constructivist approach was used to include emerging topics. Emerging codes were included during the analysis process. Except for the first transcription, codes were applied to interview transcripts using Excel by one analyst (second author). Regular meetings were held between the first and second author in order to share new codes added and discuss the findings. After coding, the analysts compared the responses to identify similarities and differences across the participants.

During the analysis process, we held several meetings to discuss the approach to the results and the addition of codes focused on experiences in healthcare services, including those related to discrimination, personnel treatment, experiences in record-keeping, and medical care, among others.

Additionally, a documentary review was conducted to describe the healthcare system in Peru and provide a better context for understanding our results.

### Ethics Statement

This study followed the guidelines of the Declaration of Helsinki and was approved by the CIEI-UPCH (UPCH Institutional Research Ethics Committee – CIEI-UPCH (SIDISI code 208877). At the point of recruitment, each participant was provided with an information sheet with details of the study. Signed consent was obtained for participation in the study, permission to be audio-recorded, and permission to be quoted anonymously in research outputs. The signed consent and permissions were obtained during the first quantitative component, and they were orally confirmed for this qualitative component. The participants were allowed to refuse to answer in any of the interview questions at any point of time. The audio-recordings were anonymized prior to transcription to avoid disclosure of the participants’ identity.

## RESULTS

The interviewed individuals reported using various healthcare systems. The decision to use one over the other is not solely dependent on the insurance system one has, as observed in Table 1. In fact, despite having EsSalud or SIS insurance, several interviewees mentioned using private or municipal services. This preference is precisely associated with perceived issues of poor quality, delays, or other deficiencies in the public systems.

The testimonies reveal that the doctor-patient interaction primarily determines experiences in healthcare facilities and perceptions of the quality of care received. This is perceived by users through 1) perceived empathy or differential and discriminatory treatment that may occur, and 2) the perception of efficiency generated in users, i.e., the extent to which the care provided meets the patient’s own expectations. While all these elements fall within the realm of the personal characteristics of the providers, they are also perceived as a result of insufficient human resources, working conditions (primarily salaries), and poor infrastructure.

**Figure 1:**
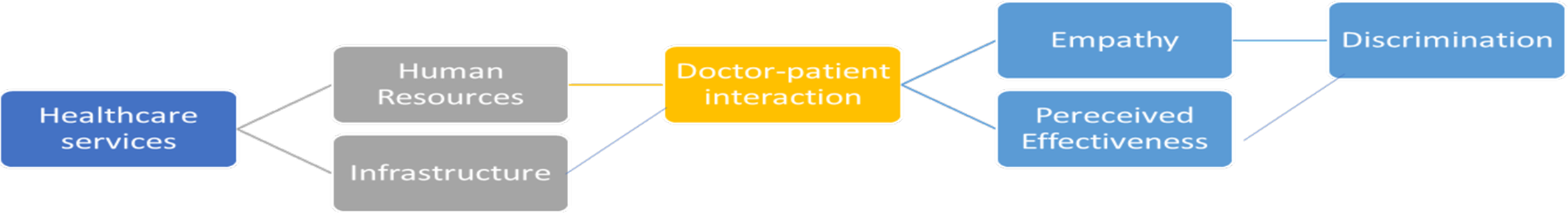
Conceptual framework of the determinants and components of the doctor-patient interaction in first-level healthcare services.

Following we will describe the three main elements that conform the perceptions of the doctor-patient interaction: the perceived effectiveness achieved by the patient, empathy, and perceived mistreatment and discrimination.

### Perceived effectiveness

Perceived effectiveness refers to the outcomes of healthcare, from the patient’s perspective. In other words, it pertains to the patient’s level of satisfaction with the care, treatment, or intervention provided. To discuss this, we can divide the care process into two key moments: The first involves the actual care itself, i.e., the moment when the patient is face-to-face with the healthcare provider and is attended to. This will be the primary moment that determines the perceived effectiveness of the patients. The second moment encompasses all the processes leading up to the care provided by the providers, including waiting times to be seen. While this is beyond the providers’ control, we address it as it will influence the perception of the overall care experience’s effectiveness.

Regarding the moment of care, we found that interviewees frequently expressed dissatisfaction with the care of their health needs in the public healthcare facilities they have visited. The shortcomings in these experiences revolve around three main aspects: 1. The perception of not receiving the medications they believe they need to alleviate their ailments, either due to shortages or because the providers did not deem it necessary to offer them; 2. The perception of not being treated adequately or receiving a clear explanation of their condition; and 3. The perception of receiving poor-quality treatments that have been poorly executed or incomplete.

(13) Women 61-65 years old Essalud user

Well, you see, I had fallen at my house, and I went to the insurance company to the polyclinic. The doctor there prescribed me three injections. She said, ‘If the pain doesn’t go away, you should go to the hospital.’ The pain didn’t go away, and after a week,I went to Hospital. I went there at the end of November, and they told me, ‘You have a problem in your hip, you need to stay for surgery.’ At Rebagliati, they have never given me therapy, even though The Dr. prescribed it.

I’ve never received therapy from either of them, or both were done over the PHONE.How can they provide post-operative therapy over thephone?

(15) Women 46-50 years old user of Essalud user

Interviewer: “Okay. And you mentioned that currently, you prefer to use that municipal or private service. Why do you prefer that service?”

Participant: “Because I will know what I have, because, well, I’ll go buy the medicines, but I’ll know what to take. On the other side, I can go to the emergency room; they give me painkillers and all, but they don’t send me for anything. Theytell me, ‘Okay, go to your health center where you’re supposed to go.’

(8) Women 46-50 years old Essalud user

“I’ve had quite a few, I’ve had a lot, but for example, one year I went to get some dental work done. I made my appointment after waiting for so long. […] I went, I had terrible pain, and thedoctor who attended to me provided a terrible service, young woman. She removed a tooth thatneeded to be removed; it was a wisdom tooth deep down that had gone bad, and she took it out, but I don’t know if the woman was having a bad day, in a bad mood, I don’t know, because she seemed like she was prying open a cow, not dealing with my body, my skin, my face, my mouth. She made me hurt terribly, but she didn’t do the necessary cleaning. After you remove a tooth, it bleeds, and many things, and you must clean the area properly, and I went home. […] Inearly died, young lady, because I spent the day,and it started hurting more, and more, and more, and I didn’t understand why…”

Regarding the appointment scheduling process, several of the interviewed individuals reported problems associated with the lengthy and cumbersome nature of the process and the fact that it must be initiated very early in the morning (with no guarantee of securing an appointment). Additionally, the process of obtaining and scheduling appointments is also affected by errors in the referral process between services, leading to further delays and inconvenience for users.

(8) Women 46-50 years old Essalud user

(Regarding the appointment scheduling process)”For example, right now, I’m waiting, and I have to wait until the 30th, 31st, to start calling at 6 in the morning to get an appointment because I stillhave a young daughter. I have to get an appointment for her and one for me. Most of the time, I don’t find any. When I call very early, at 6 in the morning, I’m on the phone dialing, and if I’m lucky, I get one, but I almost never get an appointment for the same day. Sometimes, I get one for the next day or three or four days later.

And if I’m not lucky, well, because there are many people like me doing the same, glued to their phones, calling from very early, and then you hear, ‘No appointments available,’ ‘Try again in a few days.’ Many times, I’ve waited the whole month and still couldn’t get an appointment, there are none available because they’re already booked for some areas, at least, the more complicated ones. ‘Sorry, there are no appointments, please try again.’ Then suddenly they reopen. Sometimes, you spend two, three, four days, glued to the phone, calling, calling from early morning, hoping to see if they’ve reopened so you can get an appointment.”

(2) Women 31-35 years old SIS user

Well, to get an appointment, you must schedule it very early and call starting from 6:00 in the morning to schedule it over the phone. If there are available appointments, they’ll give you one; otherwise, you’ll get it in a month or two. It’svery difficult to get an appointment sometimes.

(7) Women 41-45 years old Essalud user

“I scheduled an appointment, it was for ophthalmology, I remember. I needed it because,well, my blood pressure was bad, I remember.

They first sent me to internal medicine, and to getan appointment with internal medicine, I had to wait for a month and a half. After that, I had the internal medicine appointment, and then they referred me to refraction […]. I went for the refraction, and only then did they give me the okay and said I needed an ophthalmologist. Afterthat, they referred me to an ophthalmologist, but there were no appointments available.”

### Empathy

Empathy is associated with “interpersonal relations attributes such as effective listening, trust, respect, confidentiality, courtesy, sympathy, understanding, responsiveness, helpfulness, compassion, and effective communication between providers and clients.” The vast majority of identified negative experiences are related to empathy. Based on the nature of the mentioned elements, we can identify three aspects of empathy in which the mentioned shortcomings fall (Mercer & Reynold, 2002, #)

1. Comprehension: Understanding the patient’s situation, perspective, and feelings (and their attached meanings).
2. Communication: Effectively communicating while confirming the understanding of what has been said.
3. Action: Acting in a way that is understanding of the patient and therapeutic.

Among the testimonies obtained, there are those that indicate having perceived a lack of genuine interest or concern on the part of healthcare providers towards users. From their point of view, this translates into extremely brief visits, where there is little interest in the users’ needs and an intention to conclude the appointment quickly.

(17) Women 41-45 years old SIS user

“For example, one day, I had a doctor, and he took, I think, three, four minutes to attend to me. Just like that, very quickly. ‘Do you feel pain rightnow?’ Take this, take this. Thank you. It doesn’t seem right to me, you know?”

(10) Women 66-70 years old Essalud user

“They don’t care. Especially if we’re older people,they might think, ‘Why bother, let them die already.’ For example, I don’t feel like I’m being treated well. In general medicine, they’re terrible.

You have to wait for them to ask you, ‘Why are you here? What’s the purpose?’ But if you already have… it’s really mistreatment they always do.”

Furthermore, the testimonies reveal a strong sense of frustration among users, not only due to the lack of attention from healthcare providers regarding their reasons for seeking care but also because the system is not designed to address people’s needs comprehensively. Users perceive that the care only focuses on addressing a specific ailment or need without providing further follow-up on the patient’s case or situation.

(8) Women 46-50 years old user of Essalud user

“Look, honestly, there’s hardly… there’s no one on staff who dedicates themselves to the concern of the patient from the community where the health center is located. Mostly, I believe that everyone comes, they do their job, their work. I haven’t had the luck, you could say, that in all this time, I’ve had a doctor or a nurse who would say,’Ma’am, yes, well, I’m going to follow up with you,’ ‘I’m going to support you with your problem,’ ‘I’m going to try to provide some solution’… no, never, never. I think mostly I’ve had to go and fight.

Really, I swear, I’ve fought a lot at that healthcenter.”

As observed, the testimony not only expresses annoyance but also a feeling of frustration towards a service that does not meet expectations, and with which users must repeatedly struggle to receive care.

Associated with this, there are testimonies that the primary concern of healthcare providers is not focused on the well-being of users but rather on their ability to pay for the service or whether they have insurance. Furthermore, there is a perception that the quality of care varies depending on whether it is a paid service or not (referring to public services where the national health insurance can be used or where services can be paid for if one does not have such insurance).

(8) Women 46-50 years old Essalud user

“They are more concerned about whether the insured person doesn’t show up, if maybe their insurance expired, I don’t know, how the person is going to pay, rather than actually attending to the person. Interviewer: So, they first check that, and if not, you have to assure them how you’re going to pay..”

#### 1. Communication

While there are almost no testimonies directly referring to healthcare providers failing to explain to users the conditions they suffer from, as well as the implications and other characteristics of the treatment they will receive, considering the numerous references to the limited time available for consultations and lack of interest shown, we can infer that communication is not occurring adequately. However, the low standards that the population has regarding care mask these deficiencies. Thus, we only find one testimony in which it is mentioned that the lack of explanations from healthcare providers and the realization that these answers will not be found in the services led the interviewee to prefer a private service, where they hope to receive better care or, at the very least, a better explanation of their ailments.

(15) Women 46-50 years old Essalud and municipal user

“Interviewer: Okay. And you were telling me that you currently prefer to use this municipal, private service. Why do you prefer this service?

Participant: Because I will know what I have, because, well, I will buy the medicines, but I will know what to take. On the other hand, I can go tothe emergency room, they give me painkillers and all, but they don’t send me anything. They tell me ‘okay, go to your assigned health center’”

#### 2. Action

Among the testimonies, some highlight a lack of understanding by healthcare providers toward patients. As they report, in the face of delays in attending appointments (often for reasons beyond their control), healthcare providers do not have tolerance for time and simply close the consultation if the patient is not present at the scheduled time, even if their workday has not yet ended. It is worth mentioning that, even if they do not show up for the appointment, there is no communication system with the facilities that patients can use to report delays or other complications that may have prevented them from attending the consultation.

(7) Women 41-45 years old Essalud and private user

“The regular hours are from 8 in the morning until7 at night now, but if you arrive at 7 or later, the doctor may already be gone. If the patient doesn’tarrive on time, the doctor leaves before their scheduled end time. It has happened to me, I hadan appointment at 6:40, and due to work reasons, I arrived for my appointment, but the doctor was no longer there; they had left. Doctors, if they don’t have many patients, theyleave.”

### Discrimination and Differential Treatment

We found some testimonies that not only refer to a lack of empathy but also to more direct expressions of discrimination and differential treatment towards users. These can be divided into three types, according to the elements identified as the causes:

1. Mistreatment due to socio-economic factors (the most frequent)
2. Discrimination based on belonging to an ethnic group and/or cultural factors
3. Differential treatment attributed to a relationship between health personnel and a user

#### 1. Mistreatment due to socioeconomic factors

One of the typical situations in which users perceive differential treatment based on socioeconomic factors is when the attention of individuals who have paid for their consultation is prioritized. This occurs at the expense of users who attend the health center exercising their right as insured individuals (public health services provided by MINSA can be covered by SIS insurance or out-of-pocket expenses).

(6) Women 36-40 years old SIS user

“However, that’s where I had the bad experience with Dr. Gxxx because even though I had made my appointment, the doctor said, ‘I’m not going tosee any more patients.’ Until I heard a nurse say to him, ‘It’s a paying patient, doctor.’ ‘Oh, okay, letthem in, let them in.’ He said, ‘What’s wrong, we have to give them whatever is available.’”

#### 2. Discrimination based on belonging to an ethnic group and/or cultural factors

Among the testimonies, we did not find references to discriminatory expressions targeting the Afro-Peruvian population. In cases where discriminatory practices based on ethnic and/or cultural factors were reported, the main perpetrators were individuals who belonged to the non-medical staff of the establishment. At the same time, doctors were perceived as more tolerant of economic and ethnic differences. It was also recognized that this discrimination primarily affects people of Andean origin. These practices include the use of derogatory language against the affected population, as well as the low prioritization of their consultation.

(16) Male 46-50 years old SIS and municipal user

Interviewer:

Have you ever seen or witnessed or been told about a situation of unequal treatment?[…]

Participant:

Yes, I have seen it a couple of times. Well, sometimes they mistreat or simply don’t speak or try to help people from the highlands […]Interviewer: And when these inappropriate treatments have happened or you have seen them, what happened? Did people complain? […]Participant: At the time, people start to complain and say they should help the person. Interviewer: And in the case that you have seen, was it resolved, and was there an apology or how was it resolved in the end?

Participant: It was resolved due to the pressurefrom people, I believe, but that’s it.

(16) Woman 31-35 years old SIS user

“This also has a significant influence, racism, because there are nurses who are racist, and people from the highlands come, compatriots, with their little children, and they look at them badly and treat them as if they were nothing.”

#### 3. Differential treatment based on the existence of relationships between users and healthcare providers

In addition to ethnic, cultural, and socioeconomic factors, some interviewees identified cases of differential treatment among users based on apparent relationships between users and service providers, whether of a familial or friendly nature. These interactions lead to the prioritization of care for some individuals, at the expense of others waiting a long time to be attended to.

(7) Woman 41-45 years old Essalud and private user

“Once I saw a girl who was friends with the nurses, and she passed through quickly. I sawher arrive and go right in. The girl had just arrived, and they gave her the slip like they always give us. ‘Oh well, wait here.’ It’s totally different; she went in through a side entrance. She arrived, and she went in right away. Therewere people who complained.”

As mentioned previously, while most of the deficiencies mentioned so far fall within the realm of personnel and their own human characteristics, interviewees identify elements that could be associated with the occurrence of mistreatment and the lack of willingness to understand and address patients’ needs. The following are descriptions of these elements.

### Insufficiency of Human Resources (HR)

Among the interviewees, it is recognized that there is a shortage of human resources available for healthcare. From their perspective, this condition results in deficiencies in the services provided, such as excessive brevity in care delivery and the failure to address the ailments reported by users, primarily due to a lack of time to express them.

(8) Women 46-50 years old Essalud user

It’s all in one: a pediatrician, a general practitioner who also attends to internal medicine, and sometimes the doctor who is attending to generalmedicine must leave because they have patients who are hospitalized and they must check on them, and we always have to wait because of that. It’s not sufficient. For me, at least, in the years I’ve been receiving care at this health center, and I’ve had to deal with very strong emergencies, not just for myself but also for my daughters when they were little, it’s frustrating, really. I’ve had to fight many times, but I must take them there because you know we don’t always have enough money to go to a private clinic where you pay, take your children, and they will attend to you better. So, you must go to your health insurance. And it’s complicated,really, it’s quite frustrating.

Likewise, it is recognized that this lack of personnel affects not only the time and dedication that can be given to each patient but also how they are treated due to the fatigue and overwhelm experienced by the providers. Thus, not only is there a shortage of time for care, but also a lack of willingness to address the doubts and inquiries of the users.

(11) Male 61-65 years old Essalud user

In Essalud, excuse me, you go, they attend to you, and from the moment you enter, the people who attend to you are exhausted, stressed fromso many complaints, requests from people, demands that their tickets are worthless, the doctor told them to come back, that the stamp is missing, the date is not there, the signature is missing. The same people feel overwhelmed by working that way all day long. The spaces are shrinking more and more because of so many people going to get treated. The consulting rooms are not adequately supplied.

### Working Conditions: Infrastructure and Low Salaries

There is a perception that mistreatment and care failures result from low salaries in the healthcare system. Some interviewees identify inadequate labor arrangements for healthcare providers, partly explaining their lack of enthusiasm for patient care. The interviewees not only identify shortcomings in the attitude of healthcare providers but also recognize that the healthcare system is not creating suitable working conditions. Instead, it significantly contributes to the deficiencies found inpatient care.

(13) Woman 61-65 years old Private and Essalud user

Translate as faithfully as possible to English: “So,I, people there, talking to the workers, [I know] there aren’t many people working there. They should assign more people to attend to those with fractures. And on top of that, they are third-party personnel, from third parties who don’t get paid vacations, insurance, they don’t get paid anything. With what enthusiasm will those people attend if they are third-party personnel?”

On the other hand, the poor condition of healthcare facilities’ infrastructure and facilities causes discomfort to the interviewed users. The deficiencies found focus on two aspects: firstly, the poor maintenance of the infrastructure (walls with cracks or areas falling apart) is highlighted, and secondly, the inadequate maintenance of the premises. Regarding the latter, particular mention is made of cleanliness and the availability of basic supplies in restrooms, which are often lacking in toilet paper and soap. Furthermore, at least one person mentions that at some point, using the restroom incurred an additional cost despite the low quality of the facilities provided. Likewise, it is recognized that infrastructure failures lead to additional discomfort in healthcare services. Small, overcrowded spaces are acknowledged to generate a widespread sense of overwhelm among the users, which the interviewees identified as a factor that impacts the quality of care they receive.

(6) Woman 36-40 years old SIS user

“The bathroom had a cost. The bathroom cost fifty cents because the lady who cleaned the bathroom wasn’t subsidized by the ministry for her salary, and it was a bit for […] maintenance and even buying water because there was no water. It was the saddest thing, a health center without water. There was a tank that had facilities, basically for doctors to wash their handswith the help of a pitcher. There was no toilet paper, soap, like what you usually see now. Some things have improved. For example, I knowthat at least they no longer charge for the bathroom, but the infrastructure is still terrible. They had to fix a wall because the wall was collapsing due to the rains because it’s a very old construction. That construction at least dates to around 1981 or 1982, and the wall was made of red bricks without maintenance. There were some red Eternit roofs [corrugated fiber-cement sheets used for roofing] where they put some things, and so on.”

(8) Women 46-50 years old Essalud user

In my opinion, [the Health Center] is small for the number of patients there are. Because patients come to this center from various locations, they come here. And so there is almost never the capacity to attend to so many patients that we are. It’s complicated to get appointments. Even inemergencies. Sometimes you arrive in a bad state at the emergency room, and they give you aticket, and you are number thirty. You’re there fainting, waiting for your turn. So, I don’t think it has the capacity to attend to the number of patients there are.

## Discussion

Our study has explored the experiences of primary care utilization and the treatment received in healthcare services among the Afro-Peruvian population in Lima. We conclude that, in general, patients perceived mistreatment and rushed medical attention, a perceived lack of importance placed on patients’ needs, and a lack of willingness to empathize with and understand those needs. This perception of poor treatment, especially the shortcomings in meeting patients’ needs, results in a perception of low effectiveness of the services.

Our findings align with other studies that have explored physician-patient interactions in the primary care system, highlighting deficiencies in care (14) (15) (16). For example, a study conducted in Peru, involving 121 users of general outpatient clinics in a healthcare center in Lima, found that only 24.7% rated the doctor-patient communication as “efficient” (39.7% rated it as “deficient,” 35.6% as “average”). Only 16.6% expressed satisfaction with the care received. Furthermore, among those who rated the communication as “deficient” (48), 79% reported low satisfaction with the care, with only 4% reporting high satisfaction (17).

Additionally, narratives of discrimination in public primary care services were identified, primarily related to socio-economic factors. This discrimination becomes even more evident in services not involving direct payments (covered by state insurance) and perceived socio-economic differences. This aligns with studies like Soto-Becerra et al (18)., who found that the perception of mistreatment in social security healthcare services (ESSALUD) is associated with belonging to a lower wealth quintile (quintiles 1 or 2 compared to quintile 3). It also aligns with the study by Arpey et al. (19), where individuals believed that their socioeconomic status influenced the quality of healthcare they received, affecting various aspects, including treatment, access to care, and patient-provider interaction. The perception that their care was inferior to that provided to individuals with better socio-economic status frequently eroded trust in the healthcare system.

From the interviewees’ perspective, these deficiencies are not only a result of personal factors but also originate from system-related elements, such as a lack of human resources and poor working conditions, both in terms of salaries and infrastructure (poorly maintained and cramped spaces). It’s worth noting that these elements are referred to as factors that ultimately affect the treatment provided by healthcare providers, which is the main determinant of satisfaction with the care. This aligns with findings from studies such as Hannawa et al (20), which concluded that treatment and communication, as defining indicators of healthcare quality, often take precedence over structural or outcome-related aspects of care. It also aligns with the study by Alarcón-Ruiz (21), which found that the lack of human resources (staff deficiency) results in healthcare providers being overburdened and having little time for each patient.

Regarding the results related to the perception of ethnic or cultural discrimination, it should be considered that ethnic self-identification is a dynamic, non-homogeneous process. Specifically, as highlighted by Noles Cotito (22), the Afro-Peruvian population is in the process of constructing its self-identification as a political subject. Therefore, we can hypothesize that the absence of a significant number of testimonies of discrimination against the Afro-Peruvian population may be due to factors such as deficiencies in self-representation as Afro-Peruvian by the interviewees. In this sense, the results may differ from future explorations in social spaces with high levels of ethnic self-identification and discrimination against the Afro-Peruvian population. For example, the differences between our results and those of the Office of the Ombudsman (12), may be because for its report, this institution collected testimonies from users in cities other than Lima, where levels of perceived discrimination against the Afro-Peruvian population are higher than among other groups.

The main limitation of the research is that the entire studied population is in the province of Lima, a region with a higher percentage of Afro-Peruvian population. Despite this, our study is one of the first investigations in Peru that explores the perception of the Afro-Peruvian population in prioritized regions regarding physician-patient interaction in first-level public healthcare services. It allows us to identify shortcomings in the healthcare system that need to be overcome and highlights the importance of working on healthcare provider education regarding communication skills for improved patient interaction.

## Conclusions and Recommendations

Our findings provide insights for improving healthcare services in terms of quality of care and physician-patient interaction at primary care among the Afro-Peruvian population and, more broadly, for all populations. This includes empathy, good treatment by healthcare personnel, without discrimination based on socio-economic factors or ethnic group membership, and aspects of healthcare facility infrastructure.

Based on these findings, it is essential to conduct further research within the Peruvian healthcare system to evaluate the best strategies to improve healthcare provider-patient interaction skills. Future studies should explore the aspects of communication that are most important from the patient’s perspective for considering empathetic and compassionate care and what characteristics a curriculum aimed at improving these attributes in healthcare personnel should have.

## Data Availability

Box repository

https://app.box.com/s/xknefsprz4meuq04bzknmq4ydk31zubu

